# Probing epigenetic clocks as a rational markers of biological age using blood cell counts

**DOI:** 10.1101/2025.05.12.25327213

**Authors:** Thomas H. Jonkman, BIOS Consortium, Erik W. van Zwet, Bastiaan T. Heijmans

## Abstract

**Background:** Epigenetic clocks are widely applied biomarkers of biological age, but their biological underpinnings remain unclear. We previously showed that epigenetic clocks are affected by naïve and memory T cell proportions, suggesting blood cell composition as a potential driver.

**Methods:** Here, we quantify the contribution of cell counts to DNA methylation age (DNAmAge) and age acceleration (AgeAccel) estimated by six 1^st^- or 2^nd^-generation epigenetic clocks. First, we present a principal component analysis (PCA) method that is robust to collinearity of blood cell counts and show this provides biologically meaningful insights.

**Results:** Applying this approach, we find strong associations between DNAmAge and cell counts, particularly with naïve and memory T cells. In contrast, associations between AgeAccel and cell counts are weaker and, particularly for 2^nd^-generation clocks, primarily involve neutrophils. We validate these findings in an external dataset of artificial cell mixtures.

**Conclusions:** We conclude that DNAmAge and AgeAccel reflect different biological processes.

## Background

Aging is the leading risk factor for many diseases. However, due to the wide variation in health among older adults, researchers are increasingly interested in biomarkers that can differentiate biologically fit individuals from frail ones (1). One class of aging biomarkers are epigenetic clocks, which use age-related changes in DNA methylation (DNAm) to estimate calendar age and/or age-related clinical biomarkers. Over the past decade, several clocks have been developed (2–7). First-generation clocks, such as the blood clock developed by Hannum et al. (2), the multi-tissue clock developed by Horvath (3), and the blood/saliva clock developed by Zhang et al. (6) predict calendar age. Second-generation clocks, such as PhenoAge (4), GrimAge (5), and DunedinPACE (7), predict composite biomarkers comprised of age and clinical measurements that are more directly indicative of health.

In aging research, epigenetic clocks are usually applied by regressing DNA-methylation age (DNAmAge) on calendar age to obtain age-acceleration residuals (AgeAccel). AgeAccel is commonly interpreted as an indicator of “biological age” (8). Under this interpretation, a positive AgeAccel means that one’s “biological age” exceeds their chronological age, thus the person is prematurely aging. Indeed, AgeAccels as calculated by epigenetic clocks have been linked to numerous age-related health outcomes, including all-cause mortality, cardiovascular disease, cancer, and diabetes (9). Various commercial parties now sell kits to measure DNAmAge, claiming, for instance, that it is “the age that matters” (10) or one’s “real age” (11).

Despite the widespread application of epigenetic clocks, the concept of “biological age” remains ill-defined, and the biological mechanisms driving these clocks are largely unknown. We previously reported that first-generation clocks detect age-related changes in the proportions of naïve and activated lymphocytes in blood, with predicted ages varying by up to 40 years between purified naïve and activated T cells and NK cells from the same donor (12). However, a specific and quantitative analysis of the contribution of cell proportions to DNAmAge and AgeAccel from 1^st^ and 2^nd^ generation clocks is currently lacking.

Recent advances in cell deconvolution algorithms, such as EpiDISH (13) and IDOL-extended (14), enable accurate estimation of high-resolution cell proportions in blood samples from DNAm-data, which can subsequently be tested for associations with an outcome of interest (13). However, the analysis of cell counts is challenging due to collinearity between individual fractions, meaning that cell proportions cannot change independently. In addition to the biological correlations between fractions, they are also mathematically constrained to sum to 100%. As one cell fraction increases, another must decrease to maintain this total, making it impossible to estimate the effects of changes in individual cell fractions and health outcomes. Novel approaches are needed to address the issue of collinearity.

### We have analyzed the link between clocks and cell counts

In this study, we first outline the challenges collinearity poses when analyzing cell fractions and introduce a principal component analysis (PCA) approach to address this. We then apply this method to a dataset of 4058 individuals to investigate the associations of cell fractions with six epigenetic clocks, including 1^st^ and 2^nd^ generation clocks, both DNAmAge and AgeAccel. We find striking differences between DNAmAge and AgeAccel in both the strength of their associations with cell composition and the specific cell fractions involved. We validate these results in an external dataset of *in silico* generated artificial cell mixtures. We conclude that DNAmAge and AgeAccel represent biologically distinct phenomena, at least from the perspective of cell composition.

## Results

### Dataset and cell counts

Our dataset comprised 6 Dutch cohorts participating in the Dutch BIOS Consortium (Supplementary Table 1) for which DNA-methylation (DNAm) was measured using the Illumina 450K-array. We analyzed 4058 whole blood samples across an age range of 18-87 (Figure 1A). We used EpiDISH to estimate the proportions of 12 blood cell types (Figure 1B): neutrophils, eosinophils, basophils, monocytes, naïve and memory subsets of B-cells, CD4 T cells, and CD8 T cells, regulatory T cells (Tregs), and natural killer cells (NK cells). Neutrophils were the largest fraction, making up 56.8% of the total blood composition on average (Figure 1B). Exploratory analysis revealed age correlations for most fractions (Figure 1C). For example, naïve T cells showed negative correlations with age (R_naïve CD4 T cells_ = -0.24, R_naïve CD8 T cells_ = -0.59), whereas memory T cells showed positive age correlations (R_memory CD4 T cells_ = 0.23, R_memory CD8 T cells_ = 0.17).

**Figure 1.**
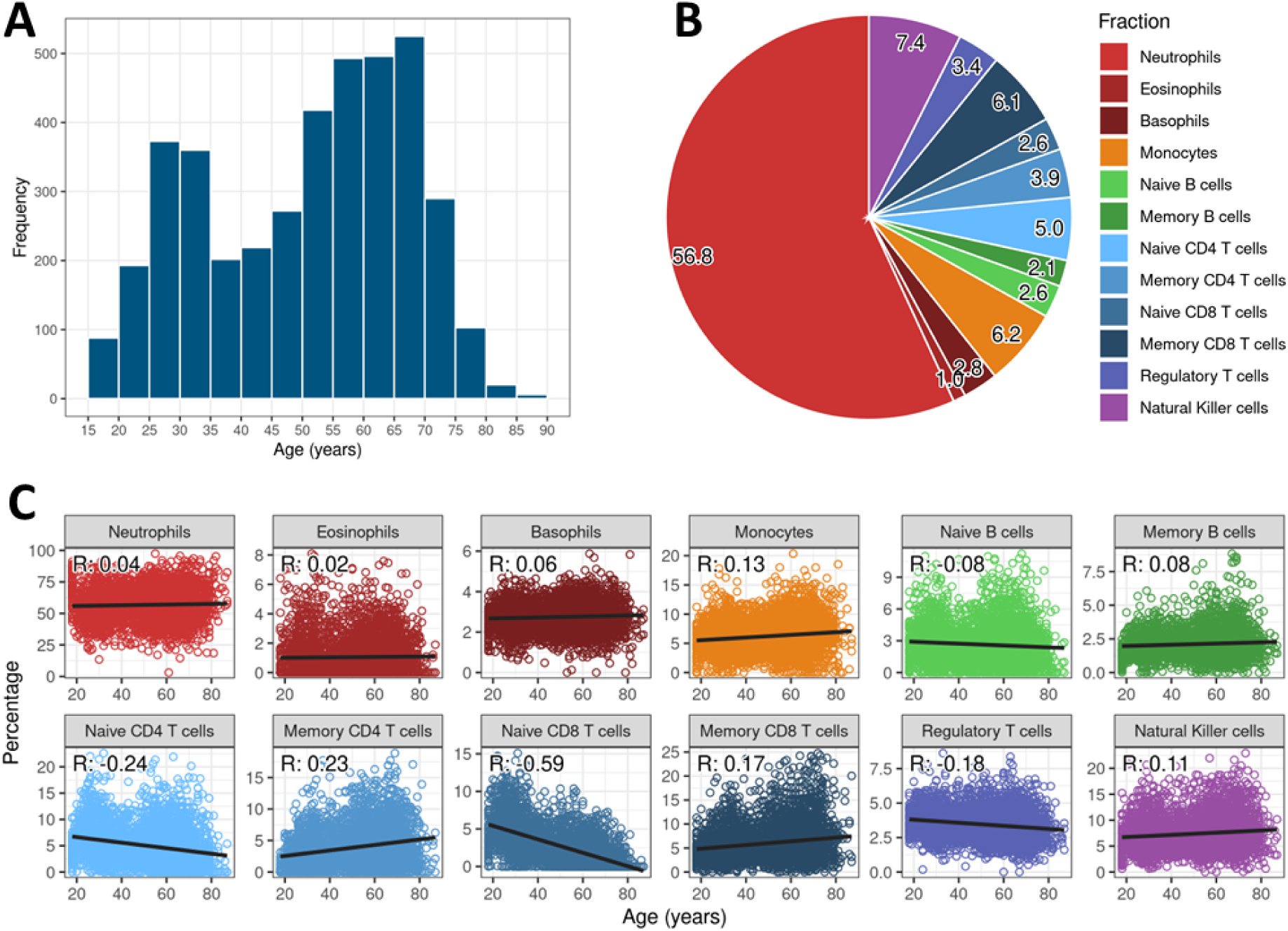
Distribution of age and cell fractions. (A) Histogram of ages of the 4058 samples included in our study. (B) Pie chart of cell fractions in all 4058 samples. The percentages of each fraction are shown on the slices of the chart. Percentages must always add up to 100% for any given person. (C) Scatter plots of percentage per cell fraction against age. Pearson correlation (R) are shown for each fraction. Trendlines represent a linear model with the fraction as the outcome and calendar age as the determinant.

### Analysis of cell fractions is complicated by collinearity

Before considering epigenetic clocks, we explored the effects of collinearity by investigating the link between cell fractions and calendar age using a univariable regression model for each fraction.

Significant associations with age (P_Bonferroni_ < 0.05) were found for all fractions except neutrophils and eosinophils (Figure 2A). Strong positive associations were observed for basophils, memory B cells, and memory CD4 T cells (effect size = 1.4, 1.4, and 1.2 years/%, respectively), while naïve CD8 T cells, regulatory T cells, and naïve CD4 T cells showed the strongest negative associations (effect size = - 3.9, -2.8, and -1.1 years/%, respectively).

**Figure 2.**
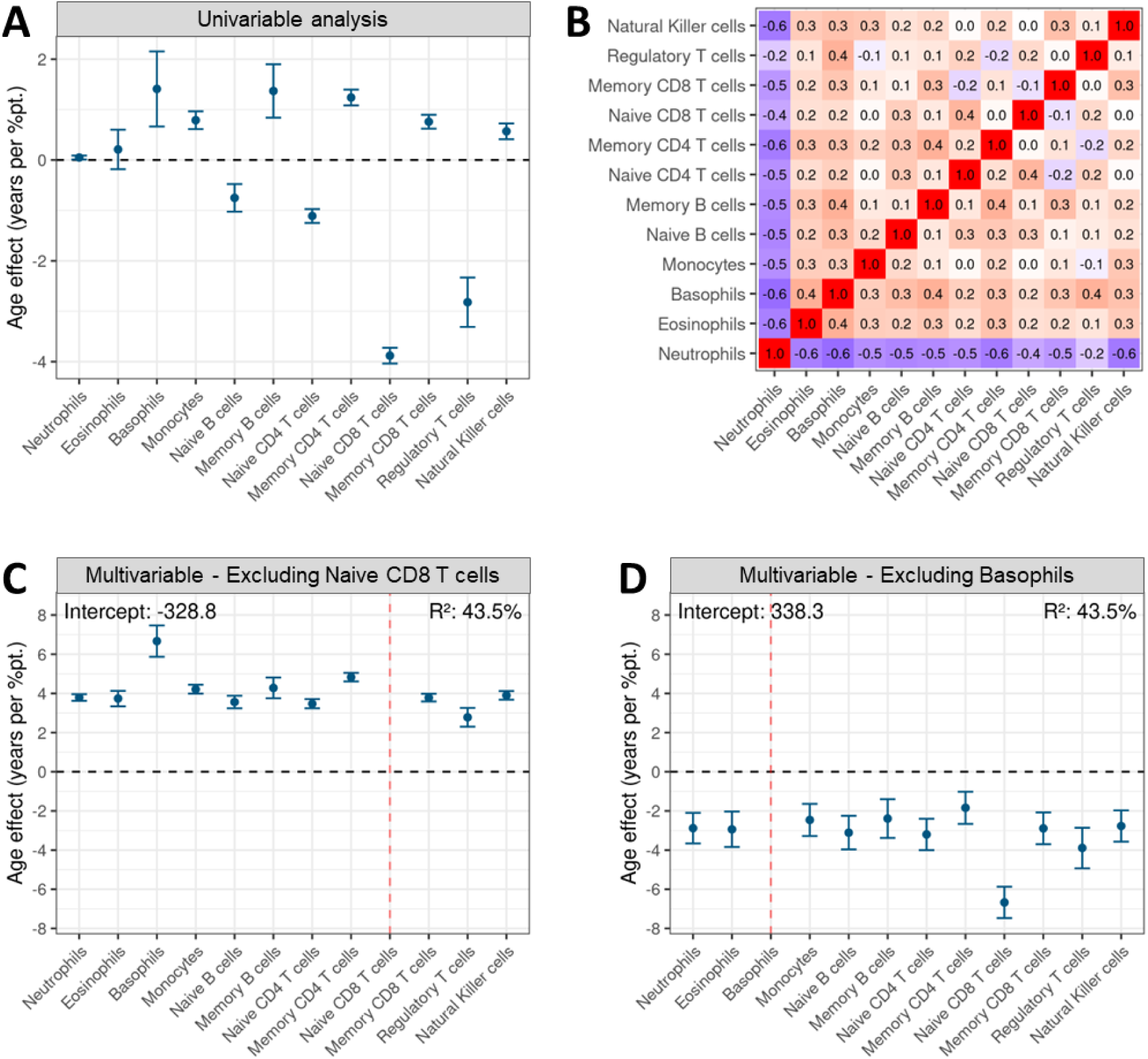
Collinearity complicates univariable or multivariable analysis of individual cell fractions. (A) Effects of 12 blood cell fractions on age from the univariable analysis. Each fraction’s effect on age was estimated independently, without considering other fractions. Effect sizes are interpreted as: "for every percentage point increase in the fraction, age increases by X years." (B) Pearson correlations between the cell fractions. (C) and (D) Effects of the 12 blood cell fractions on age from the multivariable analysis including all fractions as predictors. Due to collinearity, one fraction must be left out, skewing the effect sizes of the remaining fractions. Panel C shows results excluding naïve CD8 T cells, and panel D shows results excluding basophils. The intercepts and the percentage of age variance explained by each model are shown at the top of each panel.

However, cell fractions are correlated, both due to biological associations and because they mathematically sum to 100% (Figure 2B). Neutrophils were weakly to moderately negatively correlated to all other fractions (R ranging from -0.2 to -0.6), while the remaining fractions generally showed weak positive correlations (R up to 0.3). To assess how these correlations impacted the interpretation of effect sizes, we repeated the regression of the neutrophil fraction on age 11 times, adjusting for one of the 11 other fractions each time (Supplementary Figure 1). The effect of the neutrophil fraction on age ranged from 0.05 (no adjustment) to 0.4 (adjusting for memory CD4 T cells) to -0.3 (adjusting for naïve CD8 T cells), illustrating how collinearity makes effect sizes ambiguous.

To account for correlations between fractions, we performed multivariable analysis by including all 12 fractions in a single model (Figure 2C-D). However, since the 12 fractions add up to 100%, the 12^th^ fraction contributes no independent information (it is equal to 100% minus fractions 1 through 11), thus its effect on age cannot be estimated (shown as dashed red lines in Figure 2C-D). This 12^th^ fraction is incorporated into the model’s intercept, skewing the effect sizes for the remaining fractions. For instance, if naïve CD8 T cells are integrated into the intercept, all other fractions show strong positive age associations (P_Bonferroni_ < 10^-10^ for all fractions, Figure 2C). Conversely, if basophils are incorporated into the intercept, all other fractions show negative age associations (P_Bonferroni_ < 0.001 for all fractions, Figure 2D).

We concluded that multivariable analysis cannot produce clear effect sizes for individual fractions due to collinearity. It should be noted that this issue does not affect the total effect of all 12 fractions combined, as all models explained the same proportion of age variance (43.5%, Figure 2C-D and Supplementary Figure 1). Therefore, the multivariable approach is valid for estimating the combined effect of cell fractions, such as when adjusting an analysis for cell composition.

### PCA-based method enables collinearity-free investigation of cell composition

Principal component (PC) regression is a known approach for handling collinearity (15). PCs are linear combinations of variables—in this context, cell fractions—that are constructed to be uncorrelated. Consequently, each PC can vary independently of the others.

We calculated 11 PCs from the cell composition data (Figure 3A), corresponding to the 11 independent cell fractions. To interpret the biological meaning of these PCs, we correlated them to the individual cell fractions (Figure 3B).

**Figure 3.**
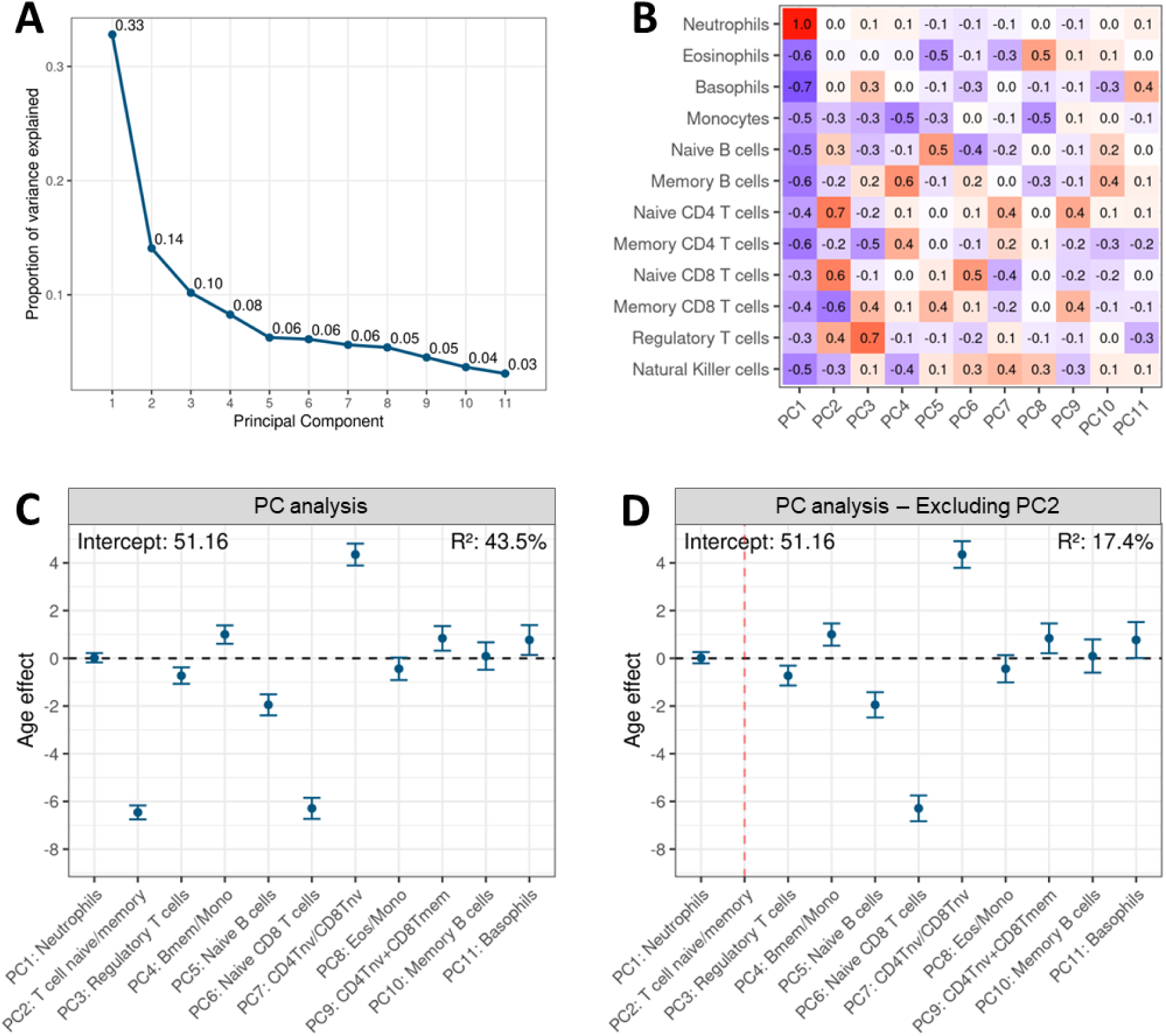
Association of PCs calculated from cell fractions with age. (A) Scree plot of the PCs calculated from cell fractions. The variance explained of each PC is shown on each bar. (B) Heatmap of correlations between the 11 PCs and the 12 cell fractions from which they were calculated. (C) Effects of the 11 PCs on age. PCs were manually labelled based on the correlations observed in Figure 3B. Effects are interpreted as: “If the PC goes up by 1 standard deviation, age goes up by X years.” (D) Effects of PCs on age when leaving PC2 out of the model. Effect sizes remain identical, with only marginal changes in the confidence intervals and a lower total explained variance.

The top PCs could reliably be interpreted as either single fractions or balances between fractions. For instance, PC1 was very strongly correlated to neutrophils (R = 1.0), and negatively to most other fractions, so we labelled it ‘Neutrophils’. This shows that the primary axis of variation in cell counts is the proportion of neutrophils relative to all other fractions. PC2 was strongly positively correlated to naïve CD4 and CD8 T cells (R = 0.7 and 0.6, respectively) and strongly negatively correlated to memory CD8 T cells (R = -0.6), thus we labelled it ‘T cell naïve/memory’. This means that the second-strongest feature in blood cell counts is the balance between naïve and memory T cell phenotypes. We labeled PCs 1-6 and PC8 in this way. For PC7 and PCs 9-11, the correlations of PCs became diffuse (no absolute correlation above 0.5); these PCs were labeled as ‘Mixed’, with an addition of their strongest correlation.

Next, we used the 11 labeled PCs in a multivariable regression model to predict age (Figure 3C). Changes in cell composition explained 43.5% of age variation (equal to the previous multivariate analysis), with the strongest associations observed for PCs representing naïve and memory T cells (PC2, PC6, and PC7, P_Bonferroni_ < 10^-10^). Specifically, PCs correlating positively with naïve CD8 T cells (PC2 and PC6) showed negative age association (effect size = -6.5 and -6.3 years/SD, respectively), while the PC correlating negatively with naïve CD8 T cells (PC7) showed positive age association (effect size = 4.4 years/SD). This aligns with the known shift from naïve to memory T cells during aging (16). Other PCs, such as those representing memory B cells vs. monocytes (PC4) and naïve B cells vs. eosinophils (PC5), also showed significant but smaller associations with age (effect size = 1.0 and -2.0 years/SD, respectively). Interestingly, while the ratio between neutrophils and all other fractions (PC1) explained a large proportion of the variance in cell composition (33%, Figure 3A), this PC was not associated with calendar age (Figure 3C), indicating that variation in the neutrophil fraction is age-independent.

To demonstrate that the PC-approach is robust against collinearity, we repeated the age regression without PC2, which had the strongest age association (Figure 3D). This pruned model’s intercept and PC effect sizes remained identical to the full model, and standard errors increased only marginally, and the total variance explained dropped to 17.4% (from 43.5%) due to the exclusion of an age-related PC. This confirmed that the PC-approach provides statistically valid and biologically meaningful results when investigating the associations of cell fractions.

### Naïve and memory T cells are strongly associated with age and DNAmAge but not AgeAccel

Next, we applied the PC-approach to study the association between cell fractions and several epigenetic clocks, considering both DNA methylation age (DNAmAge) and age acceleration residuals obtained by regressing DNAmAge on calendar age (AgeAccel). We included six clocks in our analyses, including three 1^st^-generation clocks (Hannum, Horvath, and Zhang) and three 2^nd^-generation clocks (PhenoAge, GrimAge, DunedinPACE) (Table 1). All clocks except DunedinPACE correlated very strongly with calendar age (R ≥ 0.94, Supplementary Figure 3). This was expected, since these clocks are trained to predict age (1^st^-generation clocks) or composite biomarkers composed of age and clinical measurements (2^nd^-generation clocks). DunedinPACE, which predicts the rate of longitudinal change in age-related clinical measurements rather than biological age at a single timepoint, was correlated moderately with age (R = 0.43). AgeAccel was calculated for all clocks except DunedinPACE, since the latter is already a marker of age acceleration.

**Table 1.** Epigenetic clocks included in our study. Hannum, Horvath, and Zhang are first-generation clocks, trained to predict calendar age. PhenoAge, GrimAge, and DunedinPACE are second-generation clocks, trained to predict age-related composite biomarkers. *For GrimAge, the PC-version (17) was used, as the original GrimAge clock is not publicly available. This clock is built from PCs calculated from 78464 CpGs.

| Epigenetic clock | Generation | Number of CpGs | Training samples | Training tissues | Publication date |
| --- | --- | --- | --- | --- | --- |
| Hannum | 1st | 71 | 656 | Blood | 2013 (2) |
| Horvath | 1st | 353 | 3931 | 27 tissue types | 2013 (3) |
| Zhang | 1st | 514 | 13566 | Blood, saliva | 2019 (6) |
| PhenoAge | 2nd | 513 | 456 | Blood | 2018 (4) |
| GrimAge* | 2nd | 78464 | 1731 | Blood | 2022 (17) |
| DunedinPACE | 2nd | 173 | 1037 | Blood | 2022 (7) |

First, we considered Horvath’s multi-tissue clock (Figure 4A). Since this clock’s DNAmAge correlates extremely strongly to calendar age (Supplementary Figure 3), it showed very similar associations with cell fractions. The strongest correlations were once again found for PC2, PC6, and PC7 (effect size = -6.4, -5.5, and 3.4 years/SD, respectively), which represent naïve and/or memory T cell fractions. Notably, AgeAccel associations were much weaker for these PCs (effect sizes of -0.7, 0.1, and -0.5 years/SD, respectively); PC6’s association became non-significant, and PC7’s association even reversed in direction.

**Figure 4.**
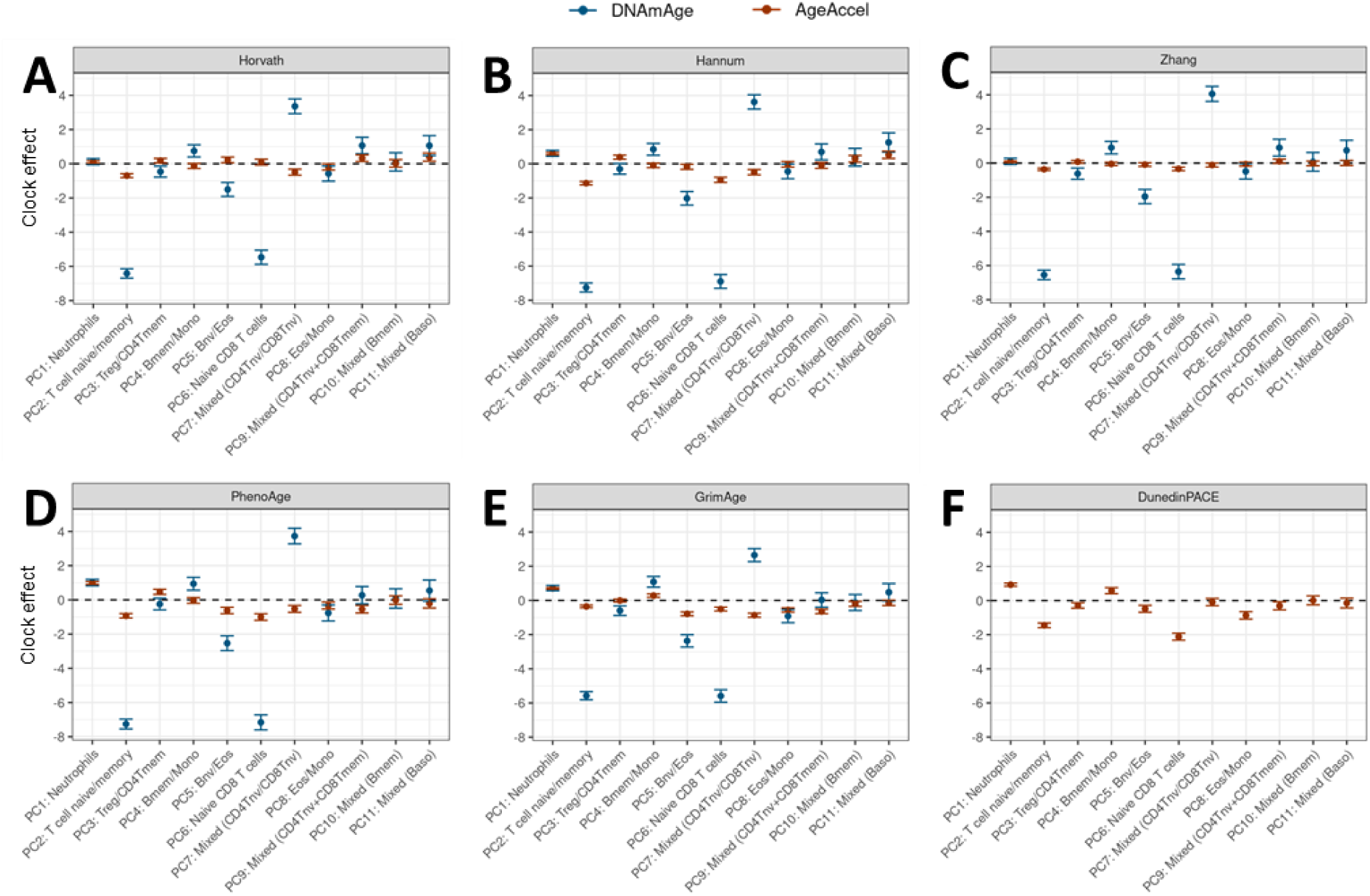
Association of cell fraction PCs with epigenetic clocks. (A-F) Effects of the 11 cell fraction PCs on the DNAmAge (blue) and AgeAccel (red) of three first-generation clocks - Horvath, Hannum, and Zhang (A-C) and three 2nd-generation clocks - PhenoAge, GrimAge, DunedinPACE (D-F). PCs were manually labelled based on the correlations observed in Figure 2B. Effect sizes are interpreted as: “If the PC goes up by 1 standard deviation, DNAmAge/AgeAccel goes up by X years.”

The six clocks included in our study have different training tissues (e.g. blood samples for Hannum and 27 different tissue types for Horvath) and predict different outcomes (calendar age for 1^st^-generation clocks and health-related composite biomarkers for 2^nd^-generation clocks). As such, the clocks could capture different biological processes and thus associate differently with cell fractions. These differences, if any, would be mostly be observed in the clock’s AgeAccels, since the DNAmAges of all clocks are strongly correlated to calendar age and thus also to each other.

When examining AgeAccel across all clocks (Figure 4B-G), we observed both shared and distinct patterns. For all clocks, AgeAccel had weaker associations with cell composition than DNAmAge, especially for the PCs representing T cell subsets (PC2, PC6, and PC7). This was especially true for Horvath, Zhang, and GrimAge (Figure 4A/C/E), where PC2 (representing T cell naïve-to-memory ratio) showed effect sizes of only -0.7, -0.4, and -0.4 years/SD, respectively. Conversely, the AgeAccel of Hannum, PhenoAge, and DunedinPACE showed stronger association with PC2 (effect sizes of -1.1, -0.9, and -1.5 years/SD, respectively).

Additionally, PC1, representing the neutrophil fraction, had relatively major effect on the AgeAccels of Hannum, PhenoAge, GrimAge, and DunedinPACE (effect size = 0.6, 1.0, 0.7, and 0.9 years/SD, respectively), but minimal effects for the AgeAccels of Horvath and Zhang (effect sizes: 0.1 and 0.1 years/SD, respectively).

In summary, we observed that the AgeAccels of Horvath and Zhang were mostly independent of cell fractions, with only minor contribution from T cell phenotypes, while GrimAge was mostly affected by the neutrophil fraction, and finally Hannum, PhenoAge, and DunedinPACE were affected by both T cells and neutrophils.

To assess the contribution of cell composition to the epigenetic clocks, we estimated the proportion of variance of DNAmAge and AgeAccel that was explained by cell composition. Cell composition explained 44% of the variance in calendar age and likewise 44 - 53% of the variance in DNAmAge for the investigated clocks (Figure 5A, panels “Age” and “DNAmAge”). Again, the primary features explaining this variance (Figure 5B, panels “Age” and “DNAmAge”) were PC2, PC6, and PC7, which all represent naïve and memory T cells. Other PCs explained only marginal proportions of the variance (≤ 2.2%).

**Figure 5.**
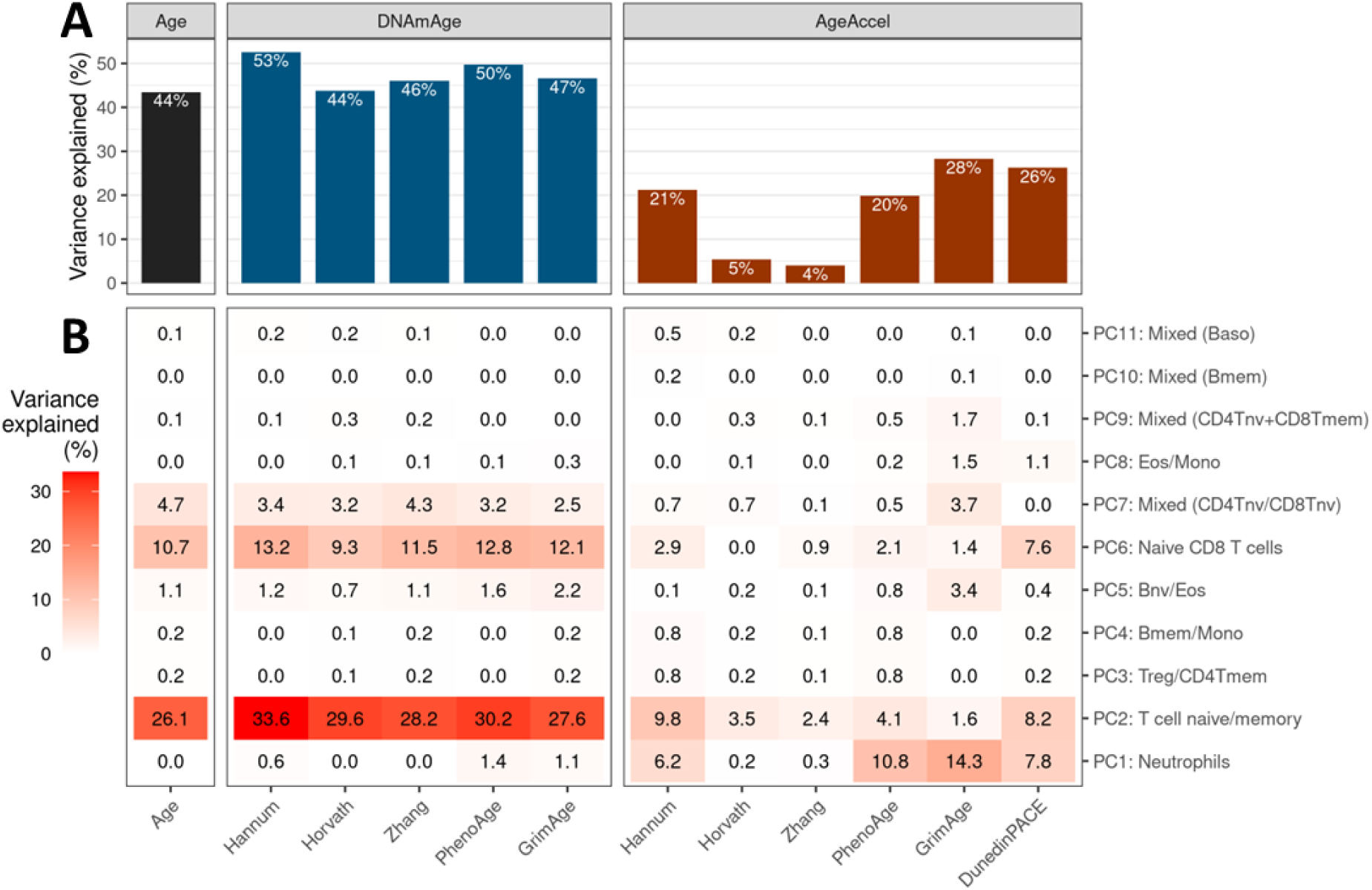
The proportion of variance of epigenetic clocks that can be explained by cell composition. (A) Total variance explained by cell composition for age (black bar), DNAmAge according all clocks except DunedinPACE (blue bars), and AgeAccel according to all clocks (red bars). Numbers show the total percentage of the given readout that can be explained by cell composition. (B) Variance explained by each individual cell composition PC. Numbers in each cell show the percentage of the given readout (columns) that can be explained by one cell composition PC (rows).

The relationship between cell composition and AgeAccel varied greatly across clocks (Figure 5A, right panel). For example, cell composition explained just 4% of the variance in Zhang’s AgeAccel, but 28% in GrimAge’s AgeAccel. This suggests that while DNAmAge behaves similarly across clocks, AgeAccel has different biological interpretations, with some being more affected by cell composition than others.

Not only the total of the variance explained by cell composition differed for the clocks’ AgeAccels, but also the contributing cell fractions (Figure 5B, panel “AgeAccel”). The AgeAccel of Hannum, PhenoAge, and DunedinPACE associated primarily with PCs representing neutrophils (PC1) and T cells (PC2, PC6, and PC7), while the AgeAccel of GrimAge associated mostly with neutrophils (PC1), with minor contribution from PCs representing T cells (PC2, PC6, PC7) and other cell types (PC5, PC8, PC9). In line with previous analyses, cell composition barely explained the AgeAccels of Horvath and Zhang, with only minor contributions from PC2 (T cell naïve-to-memory ratio) only.

The divergent associations of calendar age and AgeAccels suggest that CpGs comprising the various clocks could be consistent calendar age trackers, while others may be more informative for deviations from the norm for a given age. To explore this, we analyzed how a 1-year increase in either calendar age or AgeAccel affects the DNAm values of the CpGs used in the clocks (excluding GrimAge, which is based on PCs instead of individual CpGs; Supplementary Figure 4). We observed that the CpGs comprising first-generation clocks (Hannum, Horvath, Zhang) showed strong similarity in their effect sizes for calendar age and AgeAccel (R = 0.93, 0.88, and 0.94, respectively), but the AgeAccel effects were much less reliable than calendar age effects, with SEs being 3.5, 3.4, and 5.5 times higher, respectively (Supplementary Figure 4A-C). In contrast, CpGs making up second-generation clocks (PhenoAge and DunedinPACE) showed a lower correlation between effect sizes for calendar age and AgeAccel (R = 0.68 and -0.01 respectively), but higher reliability of the AgeAccel effect (SE ratio = 2.8 and 2.5, respectively). These results suggest that the AgeAccels of 1^st^- generation clocks contain considerable noise, while the AgeAccels of 2^nd^-generation clocks are less noisy but distinct from calendar age in terms of biological association.

Taken together, the association of DNAmAge and AgeAccel with cell fractions are strikingly different, both in magnitude and in terms of the cell fractions that contribute. Secondly, the AgeAccels of the various clocks differ in their association with cell fractions, suggesting that different clocks have different biological underpinnings.

### Cell composition drives epigenetic clocks

We observed that PCs representing neutrophils and the naïve-to-memory T cell balance were associated with AgeAccel for Hannum, PhenoAge, GrimAge, and DunedinPACE. Another approach to address collinearity is to model the effect of an increase in one fraction while proportionally reducing all other fractions. This method ignores biological correlations between fractions, but does address the fractions summing to 100%. To apply this method, we computed the average cell composition for our study (Figure 1B), and predicted the clocks’ DNAmAges for this profile as a reference. Next, we systematically created artificial cell profiles by raising one fraction by 1 standard deviation, while shrinking all other fractions proportionally to maintain the total at 100%. We then recalculated DNAmAges to analyze the AgeAccel caused by these shifts in cell composition (Figure 6A).

**Figure 6.**
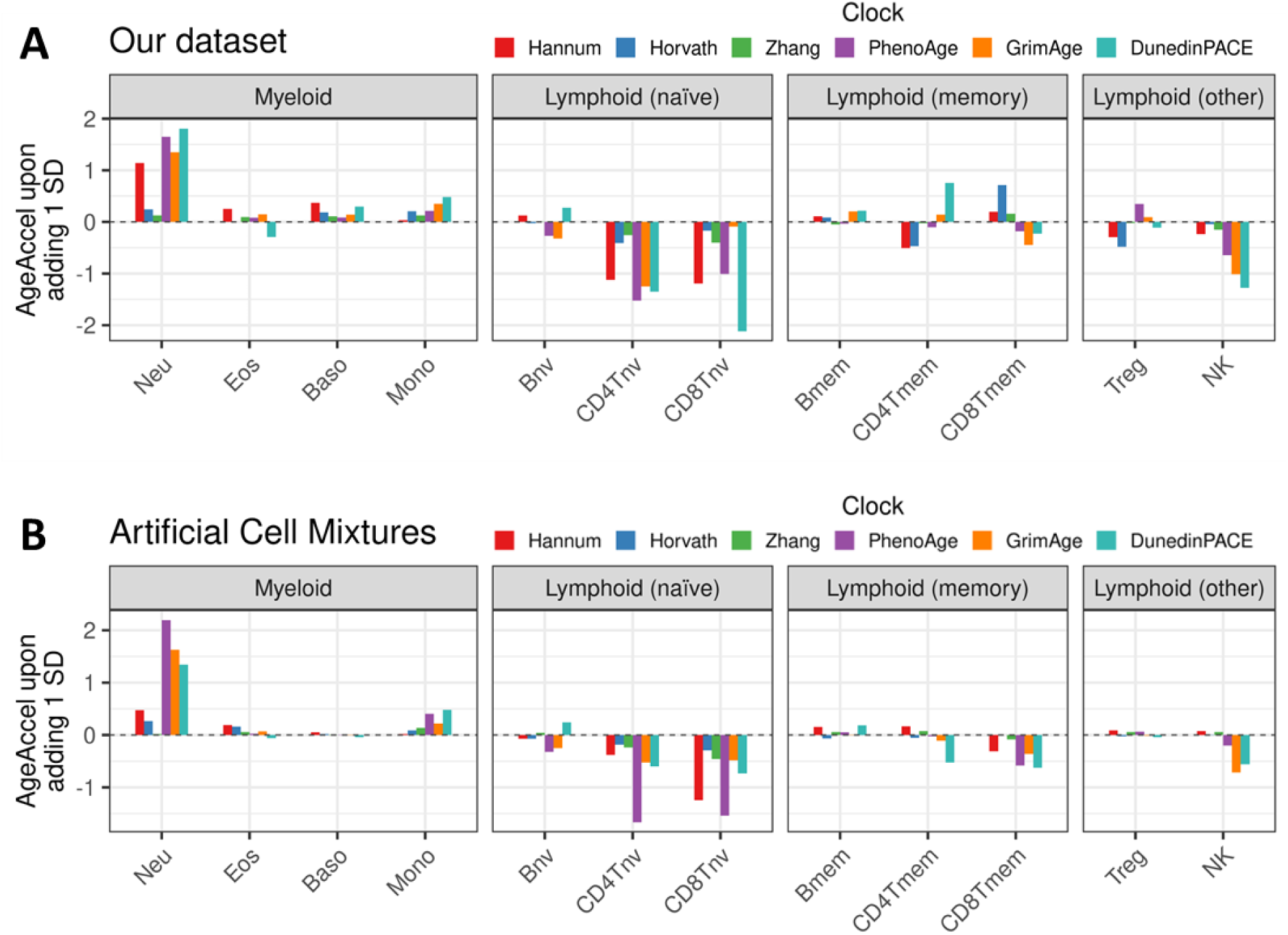
Effect of cell composition changes on AgeAccel. The change in AgeAccel observed after adding 1 SD of a cell type to a baseline sample with mean cell count distribution while proportionally shrinking all others. The X-axis shows which cell type was raised for this sample. The Y-axis shows the AgeAccel for the six clocks compared to the baseline sample (not shown, set to 0 as a reference). AgeAccels are in units of years for all clocks except DunedinPACE, for which it is in weeks. This analysis has been done in our dataset (A) and a validation dataset of in silico-generated artificial cell mixtures made from sorted blood cells downloaded from GSE167998 (B). Construction of artificial cell mixtures is described in Supplementary Figure 5.

This analysis showed that increasing naïve CD4 or CD8 T cells by 1 SD led to negative AgeAccels for Hannum, PhenoAge, and DunedinPACE (effect sizes ranging from -2.1 to -1.0 years). GrimAge showed this association for naïve CD4 T cells (effect size = -1.2 years) but not naïve CD8 T cells (effect size = -0.1 years), which may explain its weak associations with the naïve-to-memory T cells ratio (PC2, Figure 4/5). Additionally, these four clocks demonstrated positive AgeAccel in response to an increase in neutrophils (effect size = 1.1, 1.6, 1.4, and 1.8 years, respectively). Finally, we observed a negative AgeAccel for PhenoAge, GrimAge, and DunedinPACE in response to increased NK cells (effect size = -0.6, -1.0, and -1.3 years, respectively). These patterns held when repeating the analysis with naïve CD8 T cells held constant (Supplementary Figure 6), indicating that correlation between cell fractions does not substantially affect the results.

To validate these findings in an independent dataset, we downloaded an external dataset of sorted blood cell types (GSE167998), and generated *in silico* artificial cell mixtures (Supplementary Figure 5). We again started by simulating an average cell type composition, then generated additional artificial mixtures by raising one fraction by 1 SD while proportionally shrinking the others, and analyzed the effects of these changes on the clocks’ AgeAccels (Figure 6B). Despite the different datasets and methods, this analysis yielded the same effect directions as before, namely that naïve CD4 and CD8 T cells are negatively correlated with AgeAccel for Hannum, PhenoAge, GrimAge, and DunedinPACE (effect sizes ranging from -0.4 to -1.7 years), while neutrophils are positively correlated (effect size = 0.5, 2.2, 1.6, and 1.3 years, respectively). NK cells also showed mild negative effects for GrimAge and DunedinPACE (effect size = -0.7, and -0.6 years, respectively). Horvath and Zhang were once again much less driven by cell composition (effect sizes across all cell types ranging from -0.5 to 0.3 years).

We conclude that the AgeAccels of Hannum and the three 2^nd^-generation clocks evaluated (PhenoAge, GrimAge, and DunedinPACE) are indeed driven by cell composition, especially neutrophils and naïve T cells. The AgeAccels of the 1^st^-generation clocks Horvath and Zhang were much less influenced by shifts in cell composition, with only minor effects observed for naïve T cells.

## Discussion

In this study, we show how collinearity complicates the analysis of cell fractions and present a PC-based approach to address these issues. Subsequently, we use our PC-approach to show that AgeAccel has markedly different associations with cell fractions as compared with calendar age and DNAmAge. We also show that the AgeAccels of different clocks diverge in their associations with cell fractions. Finally, we show that changes in cell fractions likely drive the AgeAccel of epigenetic clocks rather than merely being correlated.

We find that calendar age and DNAmAge (irrespective of clock) are associated with a shift from naïve to memory T cell phenotypes, which is a well-known immune signature of aging (18) and in line with our previous work on first-generation clocks (12). In contrast, the AgeAccels of Hannum, PhenoAge, GrimAge, and DunedinPACE show a prominent association with neutrophils and greatly diminished association with T cells; this effect is especially pronounced for GrimAge’s AgeAccel, of which 14.3% can be explained by PC1 (representing neutrophils) and only 1.6% by PC2 (representing the naïve-to-memory T cell ratio). These findings are in line with a recent two-sample mendelian randomization study of low-resolution cell counts implying a causal effect of neutrophil and overall lymphocyte count on epigenetic clocks (19).

These findings indicate that a 1-year advancement in calendar age has different biological implications than a 1-year AgeAccel as estimated by these clocks, at least from the perspective of cell composition. These results have implications for the interpretation of epigenetic clocks as descriptors of “biological age” (20). For this interpretation to be fully valid, a 1-year advancement in calendar age would be expected to represent the same underlying biology as a 1-year AgeAccel (Figure 7). Thus, our results imply that the interpretation of AgeAccel as a direct measure of biological age may needs revision and that associations of AgeAccel with mortality and morbidity (21) may stem from processes other than biological aging.

**Figure 7.**
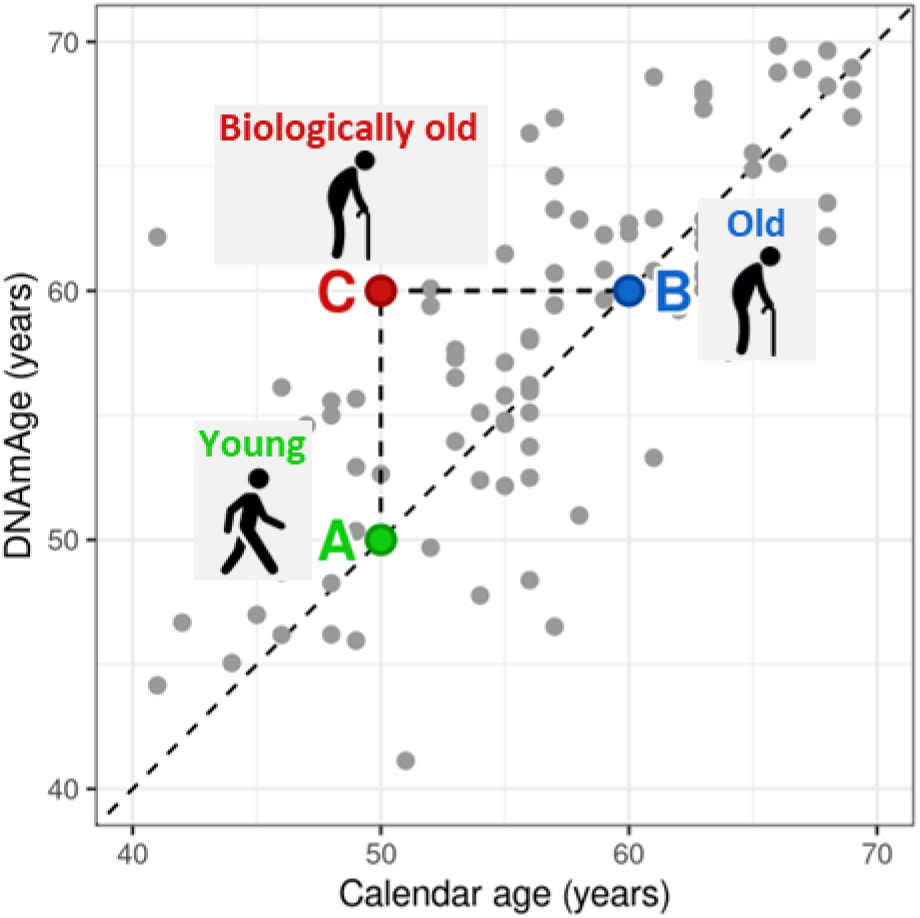
Common interpretation of AgeAccel as “biological age". This figure shows a subset of 50 participants from our dataset, with 3 people highlighted. Person A is 50 years old, while Person B is 60 years old. For both of these people, the Horvath-clock accurately estimates age. However, for person C, the clock overestimates age by 10 years. A common interpretation is that Person C is has a ‘biological age’ of 60 years, implying that they resemble a 60-year old, despite being only 50 years old. This interpretation requires DNAmAge and AgeAccel to have similar biological associations, which we do not observe in this study.

The AgeAccels of Horvath and Zhang show only very weak associations with cell composition. This could have multiple explanations. The one most commonly put forward is that these clocks’ AgeAccels capture cell-intrinsic aging information. This interpretation is supported by the fact that clocks can track calendar age even in isolated cell types (22,23) and the fact that reprogramming into induced pluripotent stem cells (iPSCs) resets the clocks to zero (24). However, since we only considered the 12 cell types included in the EpiDISH panel, the effects of other age-related immune cell types, such as effector, terminal effector, and exhausted T cell phenotypes, cannot be excluded (25). Finally, since AgeAccel is in essence an error made by an age prediction model, part of its variation must be random noise. Multiple recent studies have suggested that epigenetic clocks, particularly Zhang, are underpinned by stochastic processes (26–28). Indeed, our results indicate that the AgeAccels of Horvath and especially Zhang are noisy. The authors of the Zhang-clock previously reported that the more accurate a clock is at predicting calendar age, the less biologically relevant its AgeAccel becomes (6), posing the question if it is desirable for a biomarker of biological age to correlate strongly with calendar age. Similarly, clocks whose AgeAccel is highly affected by cell composition (Hannum, PhenoAge, GrimAge, DunedinPACE) have previously been found to have stronger associations with mortality compared to Horvath, which is less affected by cell composition (20,29). This also begs the question if a cell-intrinsic clock is desirable as an aging biomarker.

Epigenetic association studies are often adjusted for cell composition effects, but changes in cell composition could plausibly be a process through which an aging signal manifests in DNAm, for instance by tracking chronic inflammation, which is a hallmark of aging (30).

Methodologically, we analyze cell composition using a PC-based approach, since we show that univariable and multivariable models using raw cell fractions leads to ambiguous results due to collinearity. Our results suggest that methods that allow features to vary independently of one another can effectively address these challenges. We present two of such examples in this study: 1) a PC-based approach, or 2) increasing one fraction while proportionally reducing all others. A benefit of the PC-based method is that internal correlation structures between fractions are preserved; for instance, in our study, PC2 represents the ratio between naïve and memory T cell phenotypes. A limitation of this method is that PCs must be manually labeled to assign biological meaning to the results from such an analysis, which introduces a layer of complexity and subjectivity to results.

Here, we label PCs based on their correlations with individual fractions. When adopting a PC-based method, it is critical to be transparent about the PC labeling process.

We conclude that calendar age primarily associates with changes in T cell fractions while the AgeAccels of epigenetic clocks are either majorly driven by neutrophils or mostly unaffected by cell composition, depending on the clock. We expect that our results will aid in the interpretation of previous and future work in the field of epigenetic clocks.

## Methods

### Cohorts

This study was performed using DNAm data generated within the Biobank-based Integrative Omics Studies Consortium (BIOS Consortium). All data were generated in whole blood samples originating from 6 Dutch biobanks: Cohort on Diabetes and Atherosclerosis Maastricht (CODAM) (31), LifeLines (LL) (32), Leiden Longevity Study (LLS) (33), Netherlands Twin Register (NTR) (34), Rotterdam Study (RS) (35), and the Prospective ALS Study Netherlands (PAN) (36). NTR is a biobank of twins, so a random person was selected from each twin pair to ensure that samples were unrelated. We selected samples from all cohorts for which DNAm data passed quality control (QC), which was the case for 4058 samples. Calendar age was known for all samples.

### DNA methylation data

DNAm data were generated by bisulphite-converting 500 ng of genomic DNA from whole blood samples using the EZ DNA Methylation kit (Zymo Research, Irvine, CA, USA). Subsequently, bisulphite-converted DNA was hybridized onto Infinium HumanMethylation450 BeadChip arrays (Illumina, San Diego, CA, USA), and signal intensities were measured on an Illumina iScan BeadChip scanner according to the manufacturer’s protocol.

A detailed description and R code of the pre-processing workflow, including the quality control and normalization, is described in DNAmArray, which is publicly available at https://molepi.github.io/DNAmArray_workflow/. In brief, IDAT files were read into R using *minfi* (37), after which sample-level QC was performed with *MethylAid* (38). Low-quality samples were defined based on 4 QC-measures using information provided by the control probes on the array and 1 measure was based on call rate (>95%) as shown in Supplementary Figure 7, which led to the exclusion of 168 samples. Probe-level QC was based on detection p value (p < 0.01), number of beads available (≤ 2) or zero values for signal intensity. Resulting probes with more than 5% missing values were removed. 4121 samples passed QC. Normalization was done using functional normalization as implemented in *minfi*, using five principal components extracted using the control probes for normalization.

### Batch effect correction

We observed batch effects in the DNAm-data between the 6 included biobanks, therefore we performed a correction for batch effects using the *ComBat* R package (39). Age was taken as the variable of interest to prevent age differences between biobanks from being removed. To check if batch correction went correctly, PCA-plots were made of the DNAm-data before and after ComBat (Supplementary Figure 8). After ComBat, batch effects were resolved.

### Cell composition

To estimate cell composition, we used the EpiDISH deconvolution algorithm as implemented in the *EpiDISH* R package (40). We used the reference panel of 12 blood cell types (*ref.m = cent12CT450k.m*): neutrophils, eosinophils, basophils, monocytes, naïve B cells, memory B cells, naïve CD4 T cells, memory CD4 T cells, and naïve CD8 T cells, memory CD8 T cells, regulatory T cells (Tregs), and natural killer cells (NK cells). For deconvolution, we used the RPC-method (*method = "RPC"*).

Outliers in cell composition were removed as follows: for each fraction, any sample which fell at least 5 SDs away from the mean was removed. Across all samples, this led to the exclusion of 63 samples, leaving 4058 samples for analysis.

### Epigenetic clocks

Four 1^st^-generation epigenetic clocks were studied: the blood clock developed by Hannum et al. (Hannum) (2), the multi-tissue clock developed by Horvath (Horvath) (3), and the blood/saliva clock developed by Zhang et al. (Zhang) (6). Additionally, we included three 2^nd^-generation clocks, all trained on blood samples: PhenoAge, which predicts a composite score of blood health biomarkers (4), GrimAge, which predicts a composite score of smoking and several plasma proteins (5), and DunedinPACE, which predicts a composite score of longitudinal changes in functional biomarkers (7). For GrimAge, the PC-version was used (17), since the original clock is not publicly available. All clocks are described in Table 1.

Hannum, Horvath, Zhang, and PhenoAge were calculated using the *methylclock* R package (41). For GrimAge, we used the code supplied by the authors (https://github.com/MorganLevineLab/PC-Clocks). For DunedinPACE, we used the *DunedinPACE* R package (https://github.com/danbelsky/DunedinPACE).

We calculated age acceleration scores (AgeAccel) from epigenetic clocks by taking the residuals of a linear model predicting the clocks’ DNAmAge using age:

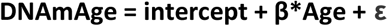

Notably, DunedinPACE is unique in the fact that instead of estimating DNAmAge, it estimates the pace of aging, i.e. the rate at which physiological function is deteriorating for a person. For this reason, we classified this clock’s estimate as AgeAccel rather than DNAmAge and did not calculate age residuals for this clock.

### Age and cell counts

A series of example analyses were performed to illustrate how collinearity between cell fractions affects models attempting to use cell fractions as predictors for an outcome, either in univariable or multivariable approaches. First, 12 univariable models were run, using age as the outcome and a single fraction as the predictor:

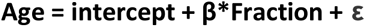

To illustrate that collinearity between cell fractions causes their effect sizes to be intertwined, we repeated this model 11 times for the neutrophil fraction, each time adjusted for one of the 11 other fractions:

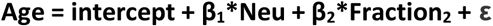

Subsequently, to demonstrate the effects of collinearity on multivariable models, 12 variations of the same multivariable models were run, using age as the outcome and all 12 fractions as predictors:

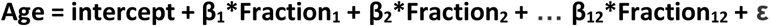

This models was repeated 12 times, each time coding a different fraction as the 12^th^. Since the effect size of 12^th^ fraction cannot be estimated due to collinearity (see Results), the 12^th^ fraction is automatically excluded from the model and incorporated into the intercept.

To solve the issue of collinearity, we calculated principal components from the cell counts. Since the cell composition contains 11 independent variables (the 12^th^ fraction being identical to 100% minus fractions 1-11), the PCA resulted in 11 PCs. PCs were labelled based on their correlations with individual cell counts to give them biological meaning (Figure 3B). We ran a model using these PCs as age predictors:

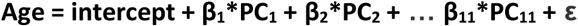

### Epigenetic clocks and cell counts

We analyzed the association between cell counts and DNAmAge/AgeAccel according to the six investigated clocks by running a linear model using the cell count PCs as predictors and DNAmAge or AgeAccel of each clock as the outcome:

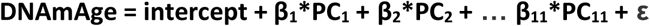

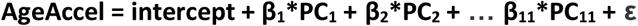

To find the variance explained by each individual PC, we ran a series of univariable models and obtained the R^2^-value:

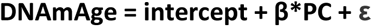

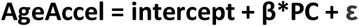

Notably, because PCs are orthogonal to each other, the effects obtained from a univariable model are identical to those from a multivariable model.

Finally, we tested the effect of raising one fraction by 1 SD while proportionally shrinking the others on AgeAccel as calculated by the clocks. To do this, we first ran a multivariable model without an intercept to model the association with AgeAccel (for each clock separately) for all cell types:

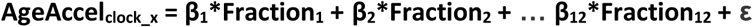

Then, we predicted AgeAccel from the average cell composition observed in Figure 1B. Additionally, we predicted AgeAccel 12 more times, each time adding 1 SD to one fraction while proportionally shrinking all others to maintain the 100% total. To obtain the effect of the cell count perturbations on AgeAccel, we subtracted the AgeAccel from the average cell composition (Figure 1B) from the AgeAccel obtained from the 12 modified cell compositions (AgeAccel_x_ - AgeAccel_avg_).

## Data Availability

The main dataset of 4058 whole blood samples is available from the European Genome-Phenome Archive (EGAC00001000277). The external dataset of 56 sorted blood cell samples is available from Gene Expression Omnibus (GEO) under accession number GSE167998.
All analyses were performed using R version 4.3.1. For all packages installed from CRAN or Bioconductor, the standard version for this R-version was used. All scripts written for this study can be found on GitHub: https://github.com/ThomasJonkmanLUMC/Cellcounts_clocks. Additionally, this repository contains integrated HTML-reports, generated by R Markdown, including both R-code and accompanying output and figures to ensure maximal transparency and reproducibility.

https://ega-archive.org/dacs/EGAC00001000277

https://www.ncbi.nlm.nih.gov/geo/query/acc.cgi?acc=GSE167998

https://github.com/ThomasJonkmanLUMC/Cellcounts_clocks

## Abbreviations

DNAmAge: DNA methylation age
AgeAccel: age acceleration
PCA: principal component analysis
PC: principal component
Treg: regulatory T cell
NK cell: natural killer cell

## Declarations

### Ethics approval and consent to participate

The study was approved by the institutional review boards of the participating centers (CODAM, Medical Ethical Committee of the Maastricht University; LL, Ethics Committee of the University Medical Centre Groningen; LLS, Ethical Committee of the Leiden University Medical Center; PAN, Institutional Review Board of the University Medical Centre Utrecht; NTR, Central Ethics Committee on Research Involving Human Subjects of the VU University Medical Centre; RS, Institutional Review Board (Medical Ethics Committee) of the Erasmus Medical Center). All participants have given written informed consent, and the experimental methods comply with the Helsinki Declaration.

### Consent for publication

Not applicable.

### Availability of data and materials

The main dataset of 4058 whole blood samples is available from the European Genome-Phenome Archive (EGAC00001000277). The external dataset of 56 sorted blood cell samples is available from Gene Expression Omnibus (GEO) under accession number GSE167998.

All analyses were performed using R version 4.3.1. For all packages installed from CRAN or Bioconductor, the standard version for this R-version was used. All scripts written for this study can be found on GitHub: https://github.com/ThomasJonkmanLUMC/Cellcounts_clocks. Additionally, this repository contains integrated HTML-reports, generated by R Markdown, including both R-code and accompanying output and figures to ensure maximal transparency and reproducibility.

### Competing interests

The authors declare that they have no competing interests.

### Funding

This research was supported by US NIH grant R01AG066887. The DNAm measurement was funded by the BBMRI-NL consortium (a research infrastructure financed by the Dutch government, NWO grants 184.021.007 and 184.033.111).

### Authors’ contributions

Conceptualization: BTH. Methodology: THJ, EWZ, BTH. Formal analysis: THJ. Resources: BIOS Consortium. Writing: THJ, EWZ, BTH. Visualization: THJ. Supervision: EWZ, BTH. All authors have read and approved the final manuscript.

## Acknowledgements

Not applicable.

## Authors’ information (optional)

Department of Biomedical Data Sciences, Leiden University Medical Center, Leiden, The Netherlands

Thomas H. Jonkman, Erik W. van Zwet, Bastiaan T. Heijmans

## Supplementary Material

**Supplementary Figure 1.**
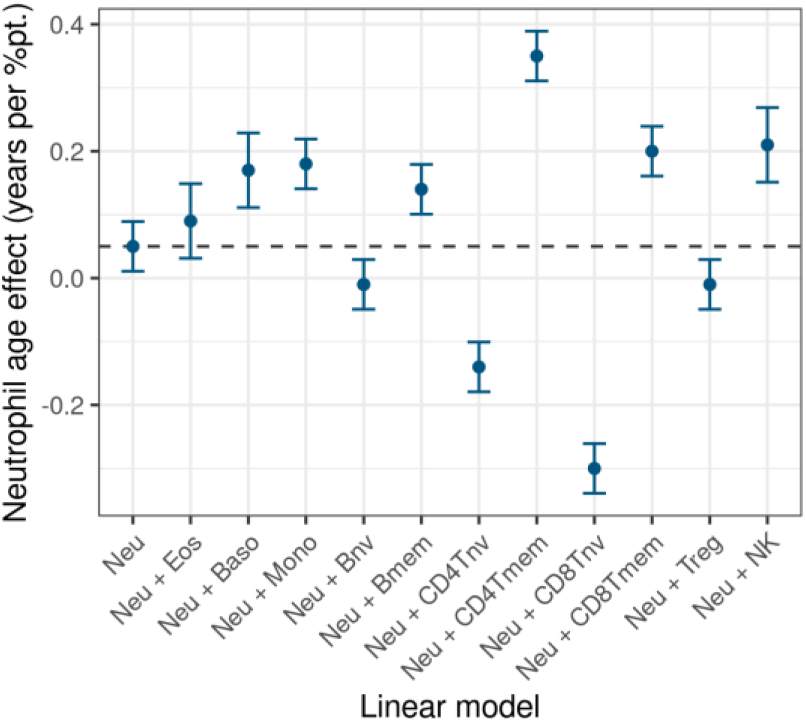
Association of the neutrophil fraction with age, with or without adjustment for one of the 11 other fractions. Y-axis shows the age effect of neutrophils according to each model (interpreted as: for each percentage point change in neutrophil percentage, one gets X years older). The model labelled “Neu” shows the unadjusted neutrophil effect, the others show the neutrophil effect adjusted for 1 other cell fraction.

**Supplementary Figure 2.**
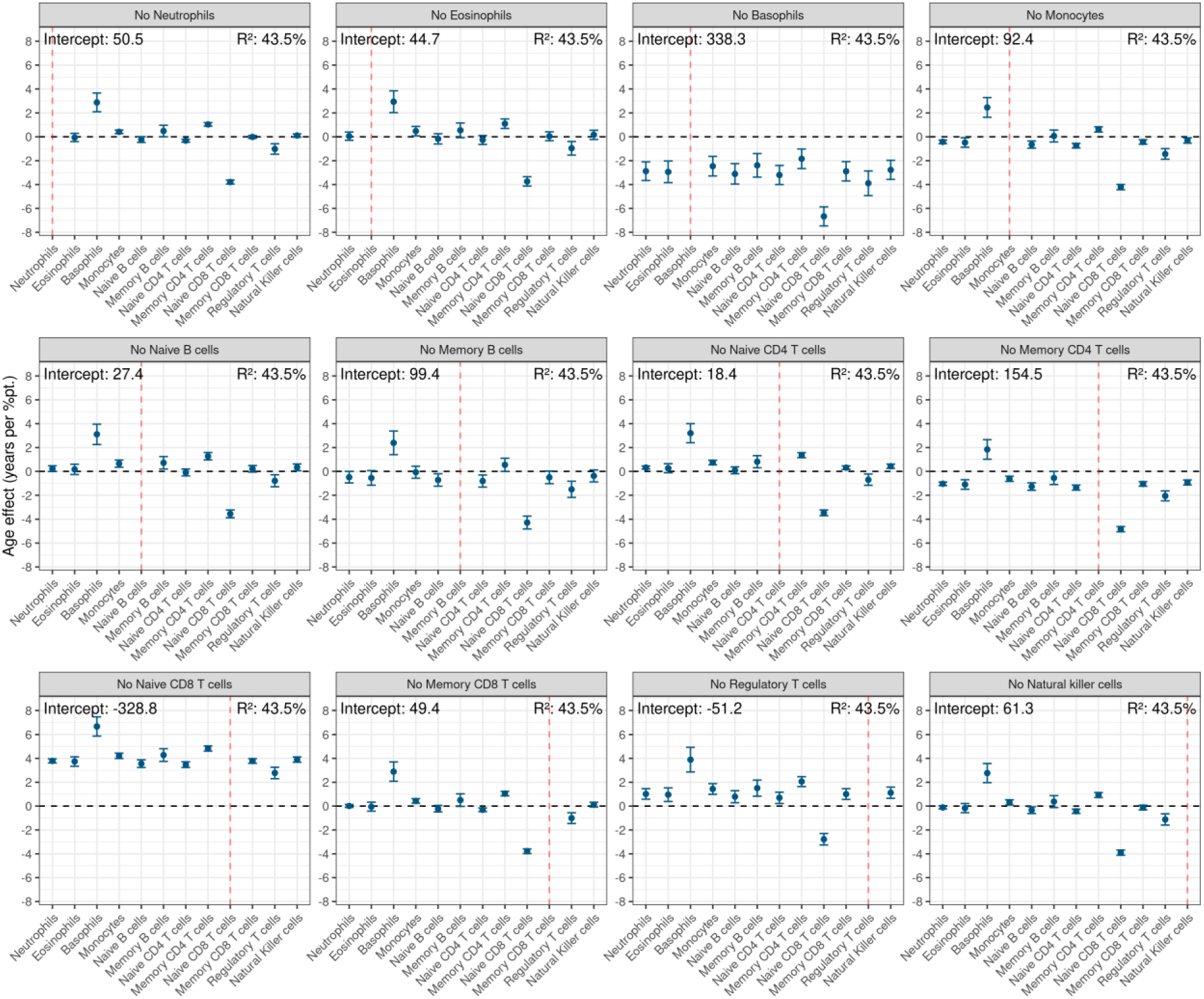
Results of all 12 multivariable analyses. The effect sizes of 12 blood cell fractions on age, from the multivariable analysis including all 12 fractions as age predictors. Collinearity forces the model to leave out one of the 12 fractions, and effect sizes of the remaining fractions are skewed by the fraction that is left out. Each panel represents the effect sizes observed in a multivariable analysis including 11 out of 12 fractions while leaving out one fraction due to collinearity. The fraction that is left out is marked with a dashed red line.

**Supplementary Figure 3.**
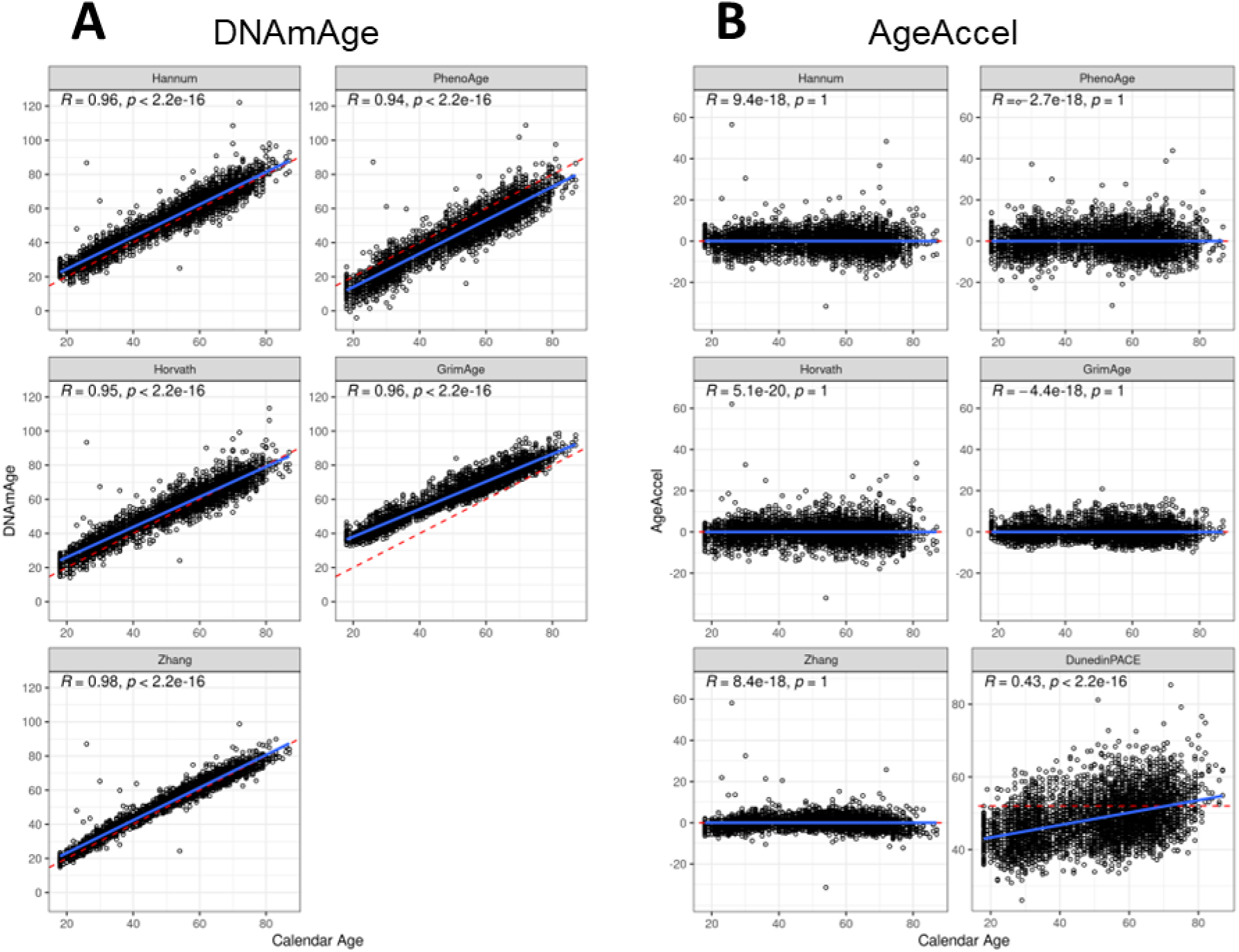
Epigenetic clocks investigated in this study. (A) DNAmAge against calendar age. Dashed red line represents the x=y line, where calendar age and DNAmAge are the same. (B) AgeAccel against calendar age, also including DunedinPACE. Dashed red line equals 0 (no AgeAccel, meaning that calendar age and DNAmAge are the same) for all clocks except DunedinPACE, where equals 52 (52 weeks of aging per year, representing a normal pace of aging).

**Supplementary Figure 4.**
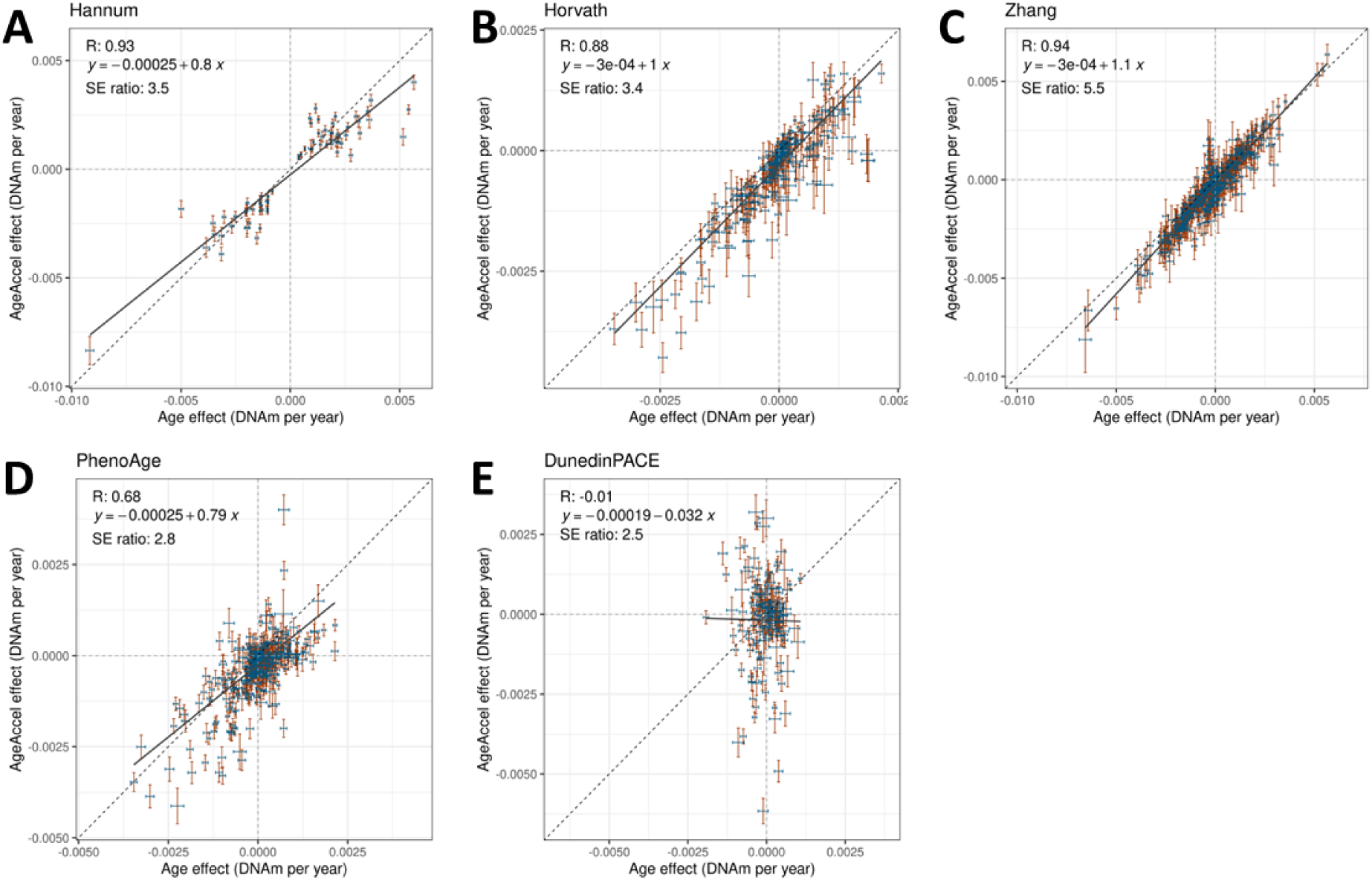
The association between calendar age and AgeAccel and DNAm of individual clock CpGs. Each panel shows the effect sizes and 95% confidence intervals (CIs) from multivariables model modelling the DNAm of each CpG comprising the clock in relation to calendar age and the AgeAccel as calculated by that clock (formula: **DNAm_CpGx_ = intercept + β_1_*Age + β_2_*AgeAccel + ε**). Each cross on the plot represents the 95% CIs of the age and AgeAccel effects of a single CpG. The calendar age effect is shown in blue on the x-axis, while the AgeAccel effect is shown in red on the y-axis. The dotted lines show the axes as well as the x=y line. Printed on each panel are the Pearson-correlation (R) between the age and AgeAccel effects, the formula of the fitted line, and the mean ratio between the standard errors (SE) of calendar age and AgeAccel (SE_Ageccel_ / SE_Age_; a higher value means that AgeAccel has a relatively high SE compared to calendar age).

**Supplementary Figure 5.**
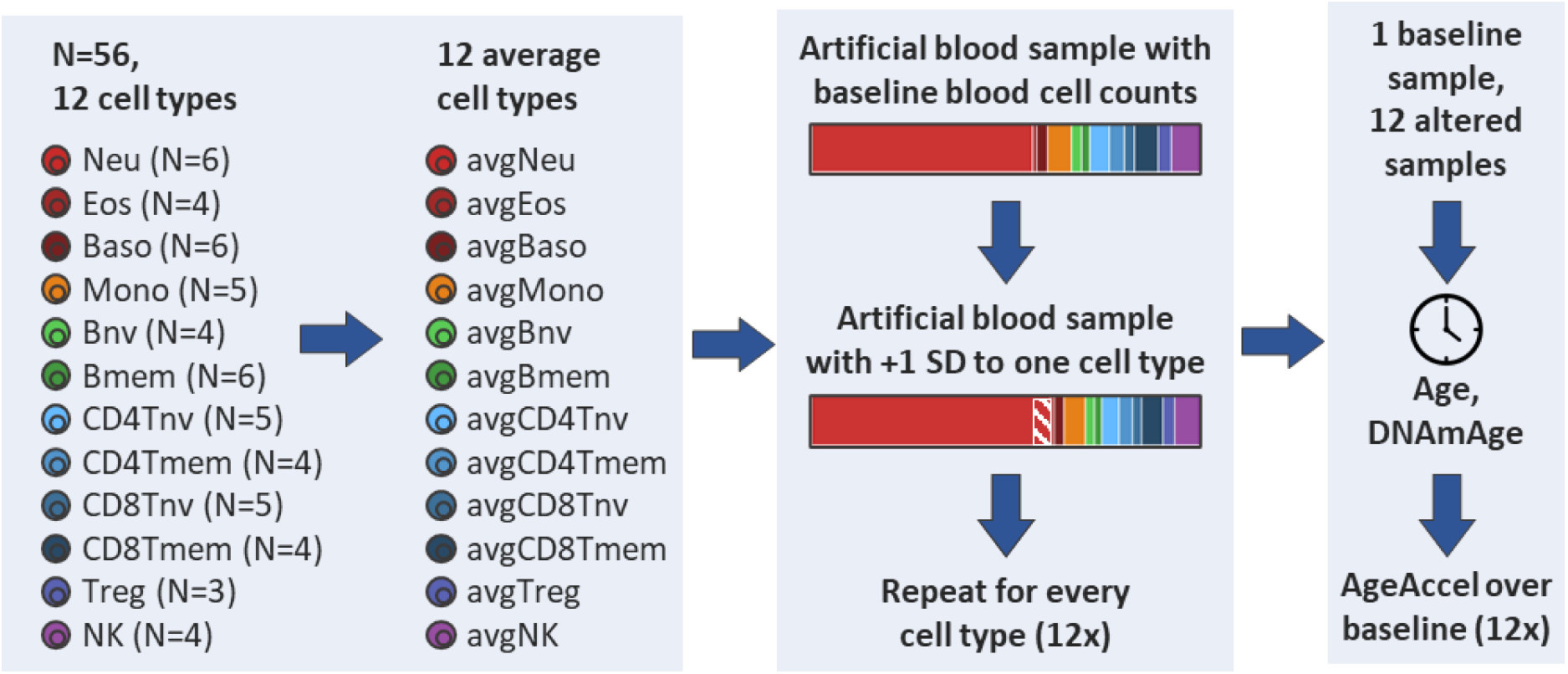
Method for constructing the artificial cell mixtures. DNAm-data from sorted cell types were downloaded from GEO (GSE167998). In total, this resource contained DNAm-data for 56 sorted blood samples. The number of samples per cell type varied from 3 to 6, and samples from different cell types originated from different donors with different ages (age range 19 - 58) We calculated average profiles per cell type by taking the mean age and DNAm-value per CpG per cell type. Subsequently, we constructed artificial cell mixtures in silico by taking the mean cell type composition found in Figure 1B (56.8% Neu, etc.). Then, we made 12 more artificial samples, each time raising the percentage of 1 cell type by 1 SD while proportionally shrinking the others to maintain the 100% total. Subsequently, we calculated a weighted mean age and DNAm for these cell type composition by multiplying the mean age and DNAm per CpG of each cell type with its percentage as shown in Figure 1B, summing these numbers for all 12 cell types, and dividing by 100%. From the DNAm-values, we calculated the clocks’ DNAmAges. AgeAccel measures were obtained by subtracting the age of the artificial sample from its DNAmAge.

**Supplementary Figure 6.**
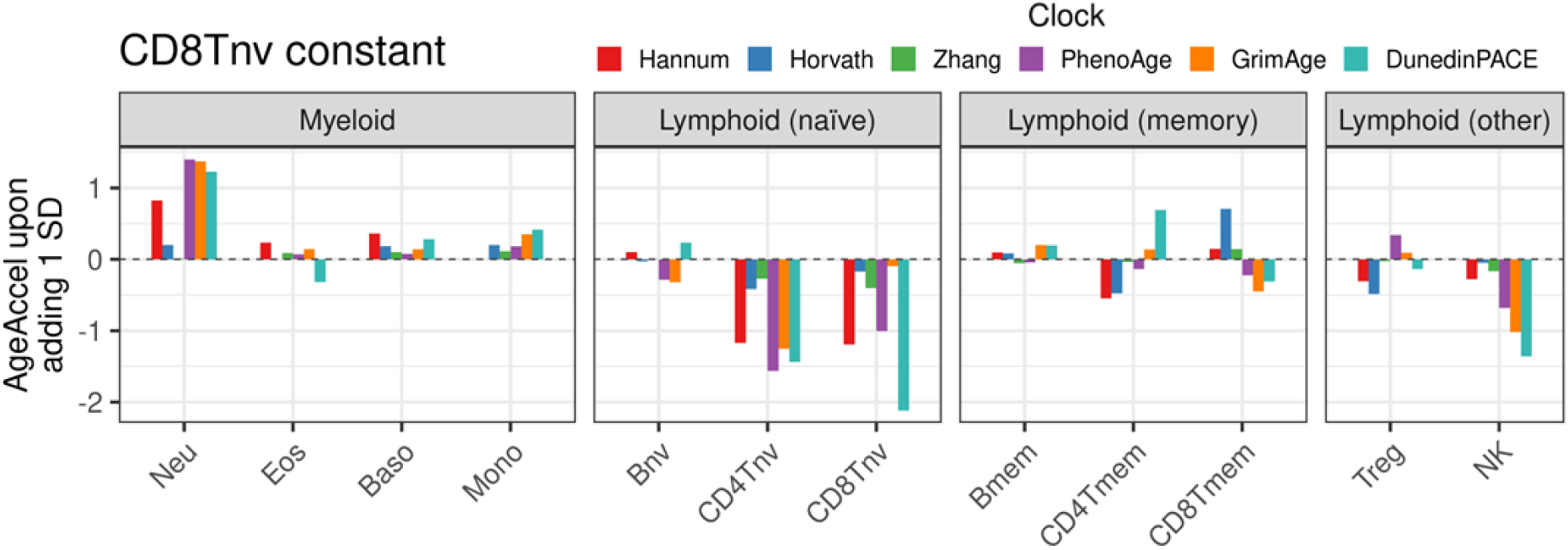
The change in AgeAccel observed after adding 1 SD of a cell type to a baseline sample with mean cell count distribution while proportionally shrinking all others except naïve CD8 T cells. The X-axis shows which cell type was raised for this sample. The Y-axis shows the AgeAccel for the six clocks compared to the baseline sample (not shown, set to 0 as a reference). AgeAccels are in units of years for all clocks except DunedinPACE, for which it is in weeks. For all fractions except CD8Tnv, all fractions except CD8Tnv were shrunk proportionally to maintain the total of 100%. For CD8Tnv, all 11 other fractions were shrunk proportionally.

**Supplementary Figure 7.**
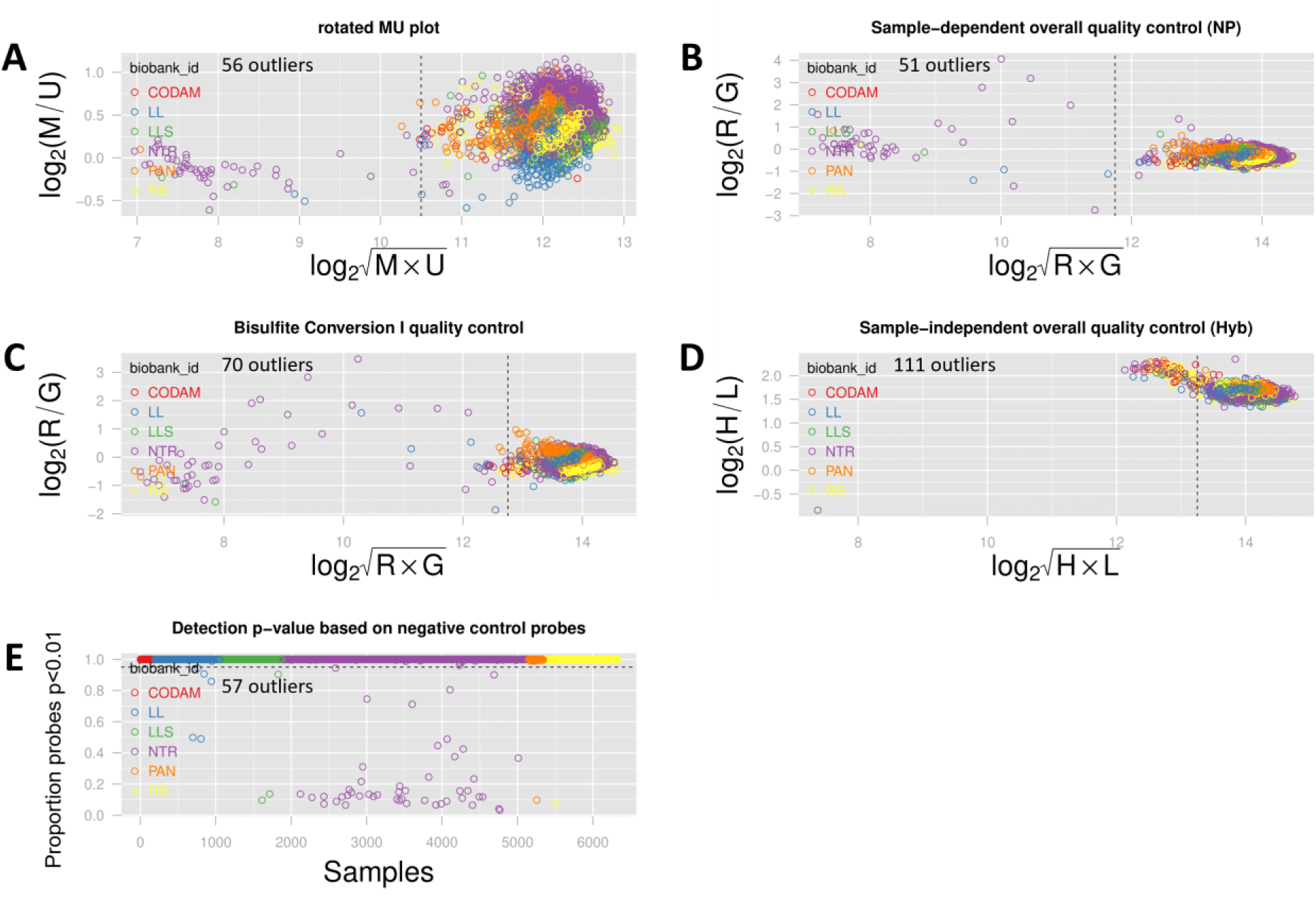
Quality-control (QC) measures for the DNAm-data. QC was based on control probe information and call rate as determined by the Methylaid software. Samples were excluded if they fell outside of at least 1 of the 5 defined QC thresholds, as shown in subfigure A-E. Samples were colored for biobank. In each panel, the number of samples falling outside of the threshold is noted. In total, 168 samples were excluded. The used cutoffs used to define outliers were as follows: (A) A median Methylated and Unmethylated log2 intensity smaller than 10.5 (rotated MU plot). (B) An average log2 intensity of the expected signals in green and red channel of non-polymorphic controls smaller than 11.75 (overall sample-dependent control plot). (C) An average log2 intensity of converted Bisulfite Type I controls in green and red channel smaller than 12.75 (bisulfite conversion control plot). (D) An average log2 intensity of High and Low hybridization controls (green channel) smaller than 13.25 (overall sample-dependent control plot). (E) Less 95% of their probes above the background signal (detection p-value plot).

**Supplementary Figure 8.**
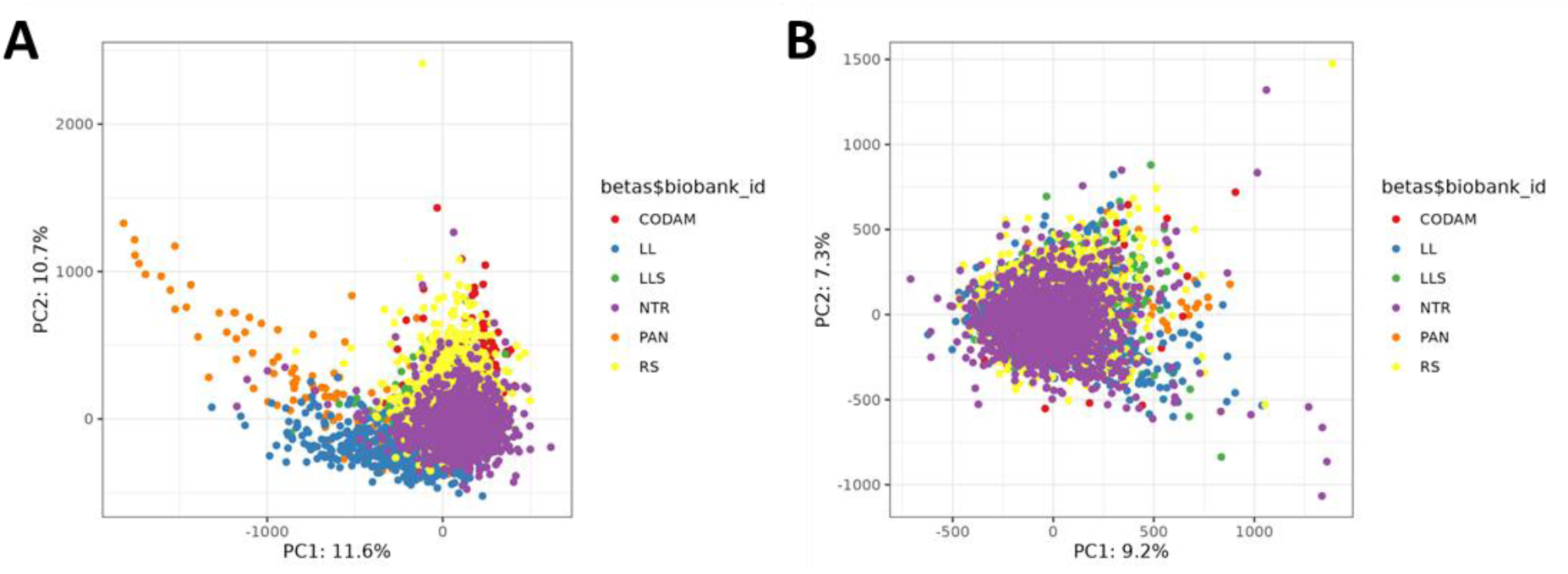
PCA of DNAm-data before and after batch correction. Dots were colored based on biobank. Before batch correction using ComBat (A), biobanks show strong batch effects, which is resolved after ComBat (B).

**Supplementary Table 1.** Cohort characteristics.

| Cohort | Number of samples | Age (years) |  |  | Male/female percentage |
| --- | --- | --- | --- | --- | --- |
|  |  | Mean | SD | Range |  |
| CODAM | 154 | 65.6 | 7.0 | 50-79 | 54%/46% |
| LL | 811 | 46.8 | 13.6 | 18-81 | 43%/57% |
| LLS | 714 | 58.3 | 6.8 | 30-79 | 48%/52% |
| NTR | 1399 | 37.6 | 13.9 | 18-79 | 34%/66% |
| PAN | 173 | 62.3 | 9.6 | 37-87 | 61%/39% |
| RS | 807 | 67.6 | 7.1 | 38-87 | 43%/57% |
| **Total** | **4058** | **51.2** | **16.4** | **18-87** | **42%/58%** |

## Notes

### Competing Interest Statement

The authors have declared no competing interest.

### Author Declarations

The Ethical Committee of the Leiden University Medical Center gave ethical approval for this work.

